# B cells, BAFF and interferons in MIS-C

**DOI:** 10.1101/2022.05.18.22275245

**Authors:** Adam Klocperk, Marketa Bloomfield, Zuzana Parackova, Ludovic Aillot, Jiri Fremuth, Lumir Sasek, Jan David, Filip Fencl, Aneta Skotnicova, Katerina Rejlova, Martin Magner, Ondrej Hrusak, Anna Sediva

## Abstract

Multisystem Inflammatory Syndrome in Children associated with COVID-19 (MIS-C) is a late complication of pediatric COVID-19, which follows weeks after original SARS-CoV-2 infection, regardless of its severity. It is characterized by hyperinflammation, neutrophilia, lymphopenia and activation of T cells with elevated IFN-γ. Observing production of autoantibodies and parallels with systemic autoimmune disorders, such as systemic lupus erythematodes (SLE), we explored B cell phenotype and serum levels of type I, II and III interferons, as well as the cytokines BAFF and APRIL in a cohort of MIS-C patients and healthy children after COVID-19.

We documented a significant elevation of IFN-γ, but not IFN-α and IFN-λ in MIS-C patients. BAFF was elevated in MIS-C patient sera and accompanied by decreased BAFFR expression on all B cell subtypes. The proportion of plasmablasts was significantly lower in patients compared to healthy post-COVID children. We noted the presence of ENA Ro60 autoantibodies in 4/35 tested MIS-C patients.

Our work shows the involvement of humoral immunity in MIS-C and hints at parallels with the pathophysiology of SLE, with autoreactive B cells driven towards autoantibody production by elevated BAFF.

**Graphical abstract:** 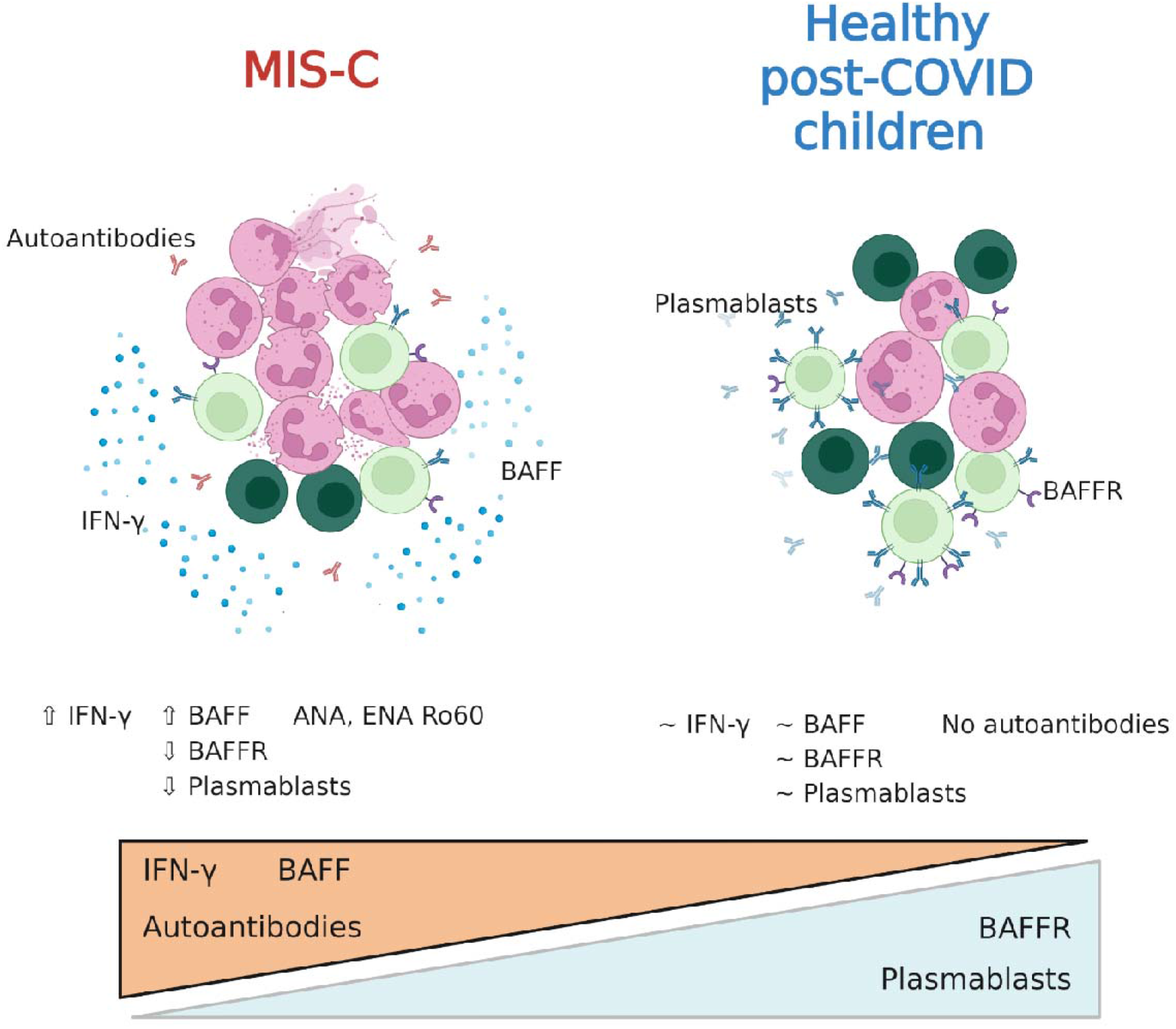

**Summary sentence:** Elevated serum BAFF in children with MIS-C supports the state of polyclonal B cell activation and autoimmune phenomena characterizing this disease.

## Introduction

Multisystem Inflammatory Syndrome in Children associated with COVID-19 (MIS-C) is now a well specified entity described in a number of excellent publications that map in detail the immune / autoimmune / inflammatory responses accompanying this condition ^1–4^. All these works point to a significant pro-inflammatory state, accompanied by alterations in cellular populations of innate and acquired immunity, with prominent lymphopenia during the acute state of the disease, which typically lags several weeks after acute SARS-CoV-2 infection, regardless of its severity. This lymphopenia is characterized by a decrease in T cells, but at the same time with their activation and clonal proliferation ^4^. Significant alterations were shown in T-cell subpopulations in MIS-C and recent reports also show their clonality and exhaustion ^5,6^.

B cells, on the other hand, are less studied in the context of MIS-C. Absolute B-cell counts were reported normal or decreased, in line with the general MIS-C-associated lymphopenia ^3,7^, however they seem to be less suppressed than T cells, with elevated proportion of B cells within the lymphocyte compartment ^4^. Some studies taking a closer look at B cell subpopulations have shown an increase in plasmablasts, as well as IgD^-^CD27^-^ double negative B cells among peripheral blood mononuclear cells ^4,8,9^. Similar IgD^-^CD27^-^ activated B cells were previously documented in systemic lupus erthematodes (SLE) in association with disease activity and autoantibody secretion ^10^. This points to a potential parallel with autoreactive B cell activation in both systemic autoimmune diseases such as SLE, and MIS-C. Consistently, a number of publications document the presence of autoimmune phenomena and autoantibodies in MIS-C patients, targeting both systemic and tissue- or organ-specific antigens^8,11^. These findings suggest a strong polyclonal antibody response driven by activated B cells. In SLE, these events are driven by the serum cytokine BAFF and APRIL ^12,13^, however, such association has not yet been studied in MIS-C.

Other shared features of SLE and MIS-C may involve interferon activation. While increased IFN-γ response has been demonstrated to be a feature shared by MIS-C and SLE ^14–16^, the activity of type I and type III interferons has only been shown in lupus ^17,18^, but not in MIS-C. Interestingly, antibodies against type I interferons have been demonstrated to contribute to COVID-19 mortality and severity ^19^.

To explore these immune factors contributing to the hyperinflammation in MIS-C we set out to assess type I, II and III interferons, serum BAFF, APRIL, and B cell phenotype and BAFFR expression in children with acute MIS-C and in healthy children after uncomplicated COVID-19.

## Patients and methods

### Patients and controls

The MIS-C cohort was recruited from patients admitted to the Department of Pediatrics, University Hospital in Motol, Prague, Department of Pediatrics, Thomayer University Hospital, Prague, and Department of Pediatrics, University Hospital in Pilsen, Pilsen, Czech Republic. Informed consent with participation in this study was signed by the participants’ legal guardians in accordance with the Declaration of Helsinki and the study was approved by the Ethical Committee of the University Hospital in Motol, reference no. EK-1376/21. Data on demographics, clinical manifestations, routine laboratory features and other investigations, therapeutic management, and outcomes were collected retrospectively from medical records of the patient, or obtained via patient/parent interview.

In MIS-C patients, samples were obtained through peripheral venepuncture after the establishment of MIS-C diagnosis, before administration of corticosteroids or immunoglobulins. Patients were included in the study based on their MIS-C diagnosis consistent with WHO criteria ^20^. In total, 50 MIS-C patients were recruited during the inclusion period between October 2020 and April 2021, 24 female, age 11 months to 18 years (7.8 ± 4.35 years, mean ± SD). The alpha (B.1.1.7) SARS-CoV-2 variant was dominant in Czechia during this period.

As a control cohort, 7 healthy children who previously underwent COVID-19, 2 female, age 1 to 14 years (9.9 ± 3.9 years), were recruited into the study (hereafter referred to as healthy post-COVID children). These healthy donors were sampled 4-6 weeks after their SARS-CoV-2 PCR positivity.

For assessment of BAFF and APRIL, 4 MIS-C patients, 2 female, age 1.4 to 5.5 years (3.5 ± 1.4 years), were re-evaluated 6 months after discharge from hospital (hereafter referred to as MIS-C convalescent). Further, 8 healthy donor children with no history of COVID-19, 5 female, age 11.6 to 17.2 years (13.7 ± 1.9 years) were included for comparison (hereafter referred to as healthy children).

Description of cohorts summarized in Table 1.

### Flow cytometry

For evaluation of peripheral blood B cell phenotype, blood was taken into EDTA-coated tubes as described above. PBMCs were obtained using Ficoll-Paque (Pharmacia, Uppsala, Sweden) and cryopreserved in liquid nitrogen.

After thawing, PBMCs were incubated in the presence of recombinant human DNAse I (Pulmozyme, Roche, Prague, Czechia; final concentration was 10 IU/mL) in complete media (RPMI 1640 supplemented with 10% of heat inactivated fetal calf serum, penicillin (50 U/mL), streptomycine (50 U/mL) and 1,7mM sodium glutamate) for 30 minutes at 37°C in a CO2 incubator. One million cells were resuspended in 100 μL PBS (Sigma-Aldrich, St. Louis, MO) and stained for 30 min in the dark at room temperature with CD5 BV421 (Cat No. 562646, BD Biosciences, San Jose, CA), IgM BV510 (Cat No. 314522, Biolegend, San Diego, CA), BAFFR BV711 (Cat No. 743573, BD Biosciences) and a dried mixture of IgD FITC, CD27 PE, CD24 PerCP-Cy5.5, CD19 PE-Cy7, CD21 APC, and CD38 APC-Cy7 (Custom-design dry reagent tube, Exbio Praha, Vestec, Czechia). Then, 2 mL of BD FACS™ Lysing Solution (BD Biosciences) were added and cells were incubated for 10 minutes in the dark, room temperature. At the end, cells were washed once in PBS with 1% BSA and pellets were resuspended in 150 μL PBS.

Flow cytometry measurement was performed on BD FACSLyrics (BD Immunocytometry Systems, San Jose, CA). FlowJo software was used for data analysis (TreeStar, Ashland, OR).

### ELISA

For evaluation of serum cytokine levels, blood was taken into uncoated tubes as described above, serum was separated by centrifugation and stored frozen at -80°C until further evaluation. BAFF and APRIL was measured according to manufacturer’s specifications using pre-made ELISA kits (BAFF from R&D Systems, Minneapolis, USA, APRIL from abcam, Cambridge, UK). Type I, II and III interferons (specifically, pan-IFN-α, IFN-γ and IFN-λ1) were quantified following manufacturer’s protocols of Human ELISA Basic KIT (HRP) (MABTECH, Sweden) using Nunc MaxiSorp flat-bottom 96-well plates (Invitrogen). Absorbance 450nm was read by multimode plate reader EnVision 2105 (PerkinElmer).

### Statistics

Statistical analysis was performed using using Brown-Forsythe and Welch one-way analysis of variance (ANOVA) and unpaired t-tests with Welch’s correction in GraphPad Prism 8.0 (San Diego, CA, USA). Values of p = 0.01-0.05 (*), p = 0.001-0.01 (**), p < 0.001 (***) and p < 0.0001 (****) were considered statistically significant.

## Results and discussion

To explore the hyperinflammatory signature of MIS-C we measured IFN-α, IFN-γ and IFN-λ in the serum of 50 patients with MIS-C sampled shortly after admission to hospital, before administration of immunosuppressive therapy, and compared them to the sera of healthy children who underwent COVID-19 4-6 weeks prior to sampling and had no signs of MIS-C. We saw no significant changes in IFN-α (t test with Welch’s correction p = 0.27) and IFN-λ levels (p = 0.33) (Fig 1A, C), therefore our data suggests that lingering type I and III interferon inflammation are not robust driving factors behind MIS-C pathogenesis, despite their role in fight against the SARS-CoV-2 infection proper ^19,21,22^. On the other hand, IFN-γ was significantly elevated in MIS-C compared to healthy post-COVID children (p = 0.0004) (Fig 1B). This elevation of IFN-γ has been described in MIS-C previously ^14,15,22^ and may reflect the concurrent T cell activation^15,22^.

**Figure 1.**
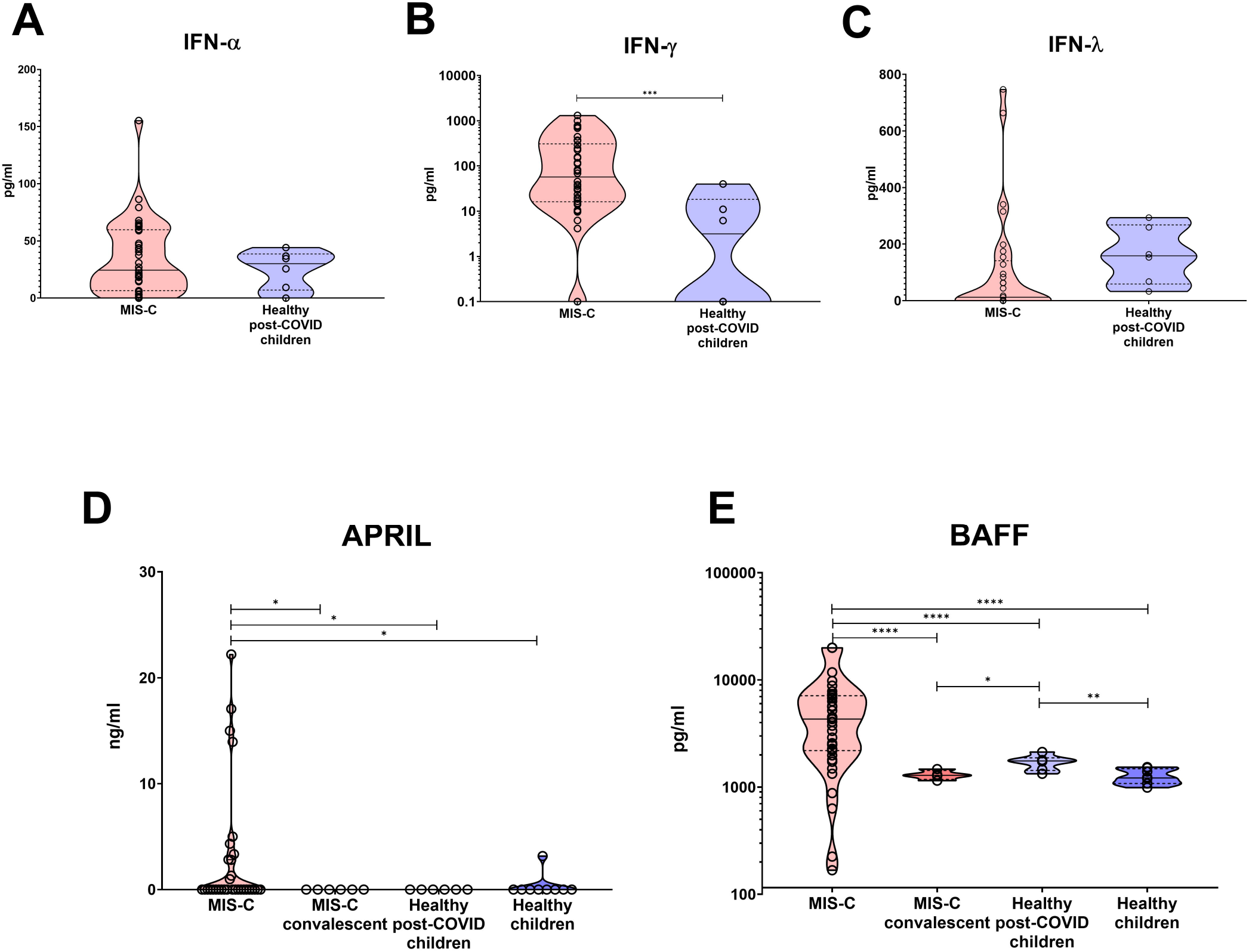
Cytokines in MIS-C patients. A, IFN-α, B, IFN-γ and C, IFN-λ in MIS-C patients and healthy post-COVID children. D, APRIL and E, BAFF in MIS-C patients, convalescent MIS-C patients 6 months after disease resolution, healthy post-COVID children and healthy donor children with no history of COVID-19 or MIS-C.

As discussed earlier, the elevation of IFN-γ and presence of autoimmune phenomena in MIS-C is reminiscent of SLE and warrants exploration of B cell immunity in these patients ^16,23^. Thus, we measured the serum concentration of BAFF and APRIL, two cytokines supporting the development and survival of B cells. The serum levels of APRIL were largely below the assay detection limit, although in a subset of MIS-C patients we detected elevated APRIL levels, which were missing in healthy post-COVID children, healthy children, and even in convalescent MIS-C patients sampled several months after full recovery (Figure 1D). Nevertheless, the majority of samples tested APRIL-negative and as such the differences were not significant.

The serum BAFF levels, on the other hand, varied significantly among the MIS-C, convalescent MIS-C, healthy post-COVID and the healthy children (Brown-Forsythe ANOVA p < 0.0001) (Figure 1E). The MIS-C patients had the highest BAFF levels of all cohorts, which in particular were higher than those in healthy post-COVID children (t test with Welch’s correction p < 0.0001), but also than those in convalescent MIS-C patients (p < 0.0001). Interestingly, even healthy post-COVID children had elevated serum BAFF levels compared to healthy children without history of COVID-19 (p = 0.0093). In both MIS-C patients and healthy children, the trend remained identical, with higher BAFF during/after disease, and lower BAFF after full recovery or in times of full health. To the best of our knowledge, BAFF has not yet been studied in connection with MIS-C and no comparison with other studies is therefore available.

The significant increase of BAFF levels in MIS-C is in line with polyclonal B cell activation, which may underlie the genesis of MIS-C-related autoantibodies and its clinical manifestations, i.e. the autoimmune systemic and organ inflammation. In our cohort, of 35 patients in whom autoantibodies were tested, 4/35 (11%) had positive Ro60 antibodies and further 4/35 (11%) had positive extractable nuclear antigen (ENA) screening but none of the tested individual antigens were positive. Previous studies in MIS-C patients reported the presence of anti-La, a characteristic autoantigen of SLE and Sjogren’s disease, and anti-Jo-1, characteristic for idiopathic inflammatory myopathies ^24^. In addition, a number of other non-routine autoantibodies directed against various autoantigens were detected, confirming the autoimmune disposition accompanying the acute stage of MIS-C ^24^. Furthermore, the combination of increased IFN-γ and a consequent increase in BAFF has been previously described in SLE, representing another analogy with MIS-C ^25^. Interestingly, in case of SLE, it has been shown that neutrophils can contribute to an increase in BAFF and augment the autoimmune process. In the case of COVID-19 and especially MIS-C, characterized by marked neutrophilia, a similar parallel might contribute to the induction of autoimmune phenomena ^26^.

Evaluating the impact of upregulated BAFF signalling on B cell subpopulations, we saw a highly significant decrease of circulating plasmablasts (p < 0.0001) and less significant decrease of transitional B cells (p = 0.029), whereas other developmental subsets did not differ significantly (Figure 2A, B). The highly significant lack of plasmablasts is of particular note given their comparatively lower reliance on BAFF for survival, which can also be bolstered by APRIL ^27^. The decrease seems to be in contrast to some previous publications ^2,4^, however, the samples analyzed here were obtained strictly before the initiation of immunomodulatory therapy (immunoglobulins or steroids), which was not always the case with the previously mentioned works. Furthermore, we calculate these subpopulations as proportion of total B cells, rather than all lymphocytes or the whole PBMC compartment, which are both disproportionately affected by the profound T cell lymphopenia characteristic for MIS-C. Indeed, our dataset also documented a relative expansion of B cells in lymphocytes (Supplementary Figure 1), which resulted in falsely comparable proportion of plasmablasts and elevated naïve B cells when calculating these subsets as proportion of all lymphocytes (Supplementary Figure 2). In the context of the absolute peripheral blood B cell lymphopenia seen in MIS-C ^3^, however, it becomes clear that plasmablasts are in fact decreased, and that not only T and B cells, but even specifically B cell subsets are differentially affected by MIS-C.

**Figure 2.**
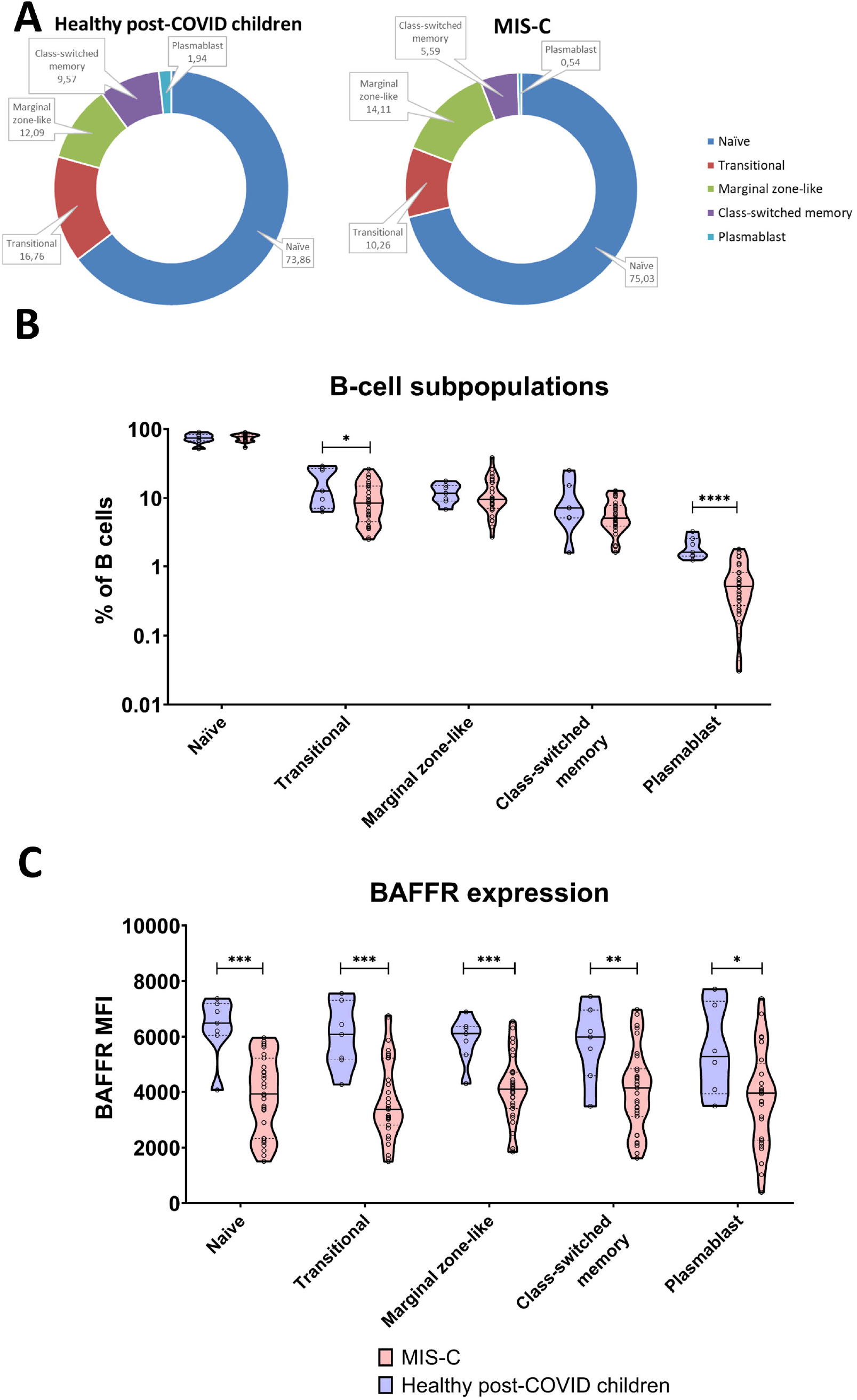
B cell phenotype in MIS-C patients. A and B, B cell subpopulations in MIS-C patients and healthy post-COVID children. C, BAFFR expression in naive, transitional, MZ-like, switched memory and plasmablast B cells of MIS-C patients and healthy post-COVID children.

Naïve and transitional B cells are also highly dependent on BAFF for survival ^28^, however all MIS-C B cells had strikingly suppressed BAFF receptor (BAFFR) expression (Figure 2C). This was true in all B-cell subsets, including naïve and transitional forms. These results are consistent with the similar situation observed in SLE and Sjögren syndrome, where high levels of BAFF are associated with decreased BAFFR expression on all B cell subtypes ^29,30^. This inverse correlation may be attributed to down-regulation of BAFFR resulting from chronic exposure to elevated BAFF levels, which was suggested to be mediated via unspecified post-transcriptional mechanisms ^30^.

In summary, our brief report highlights the dysregulation of humoral immunity with autoimmune bias in MIS-C and suggests a role of the BAFF-BAFFR axis, which warrants further exploration.

## Data Availability

All data produced in the present study are available upon reasonable request to the authors.

## Authorship

AK co-designed the study, processed samples, gathered clinical data, analysed the data and co-wrote the manuscript. MB co-designed the study, recruited patients and healthy donors, gathered clinical data, reviewed and edited the manuscript. ZP and LA performed experiments. MM, JF, LS, JD and FF recruited patients and provided clinical data. AS, KR and OH performed experiments and analysed data. AS designed the study and co-wrote the manuscript.

## Acknowledgements

The work was supported by AZV NU20-05-00320 and NU20-05-00282 issued by the Czech Health Research Council and Ministry of Health, Czech Republic and institutional support of research organization #00064203 from University Hospital in Motol, Czech Republic.

