## Supplementary Data for "B cells, BAFF and interferons in MIS-C"

Supplementary Figure 1 B cells of MIS-C patients and healthy post-COVID children shown as percentage of lymphocytes.

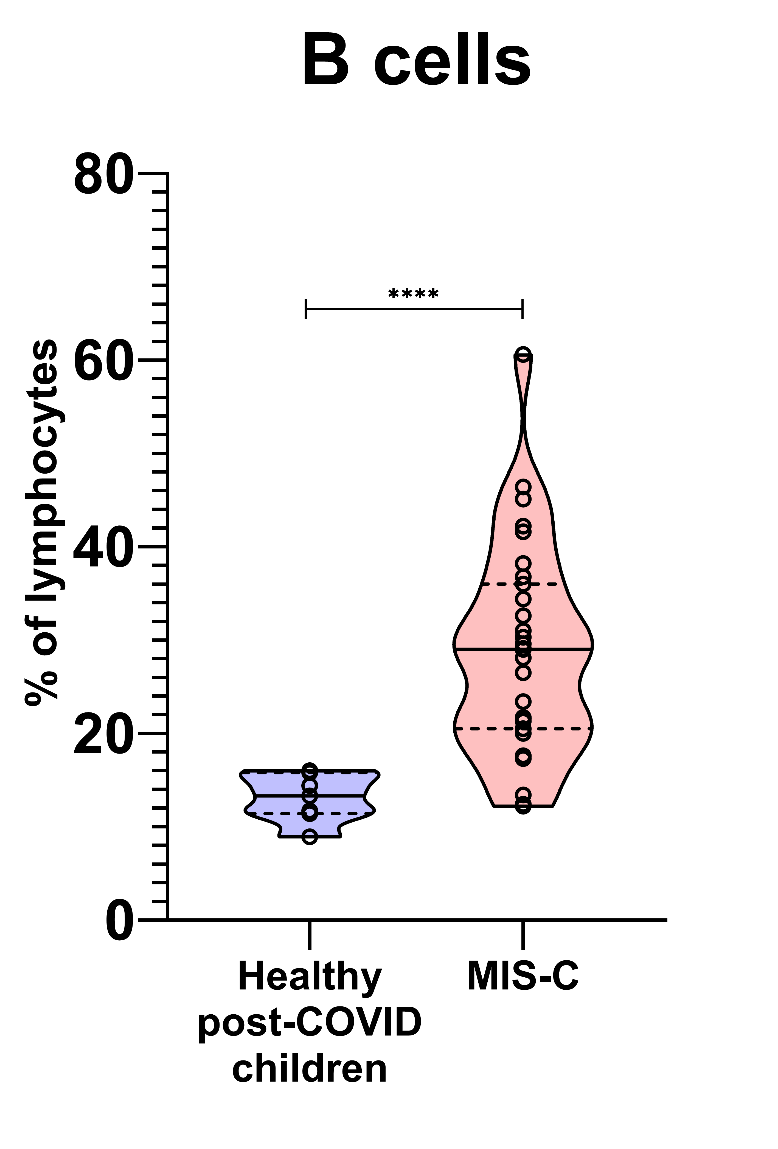

Supplementary Figure 2 B cell subpopulations in MIS-C patients and healthy post-COVID children, shown as percentage of lymphocytes.

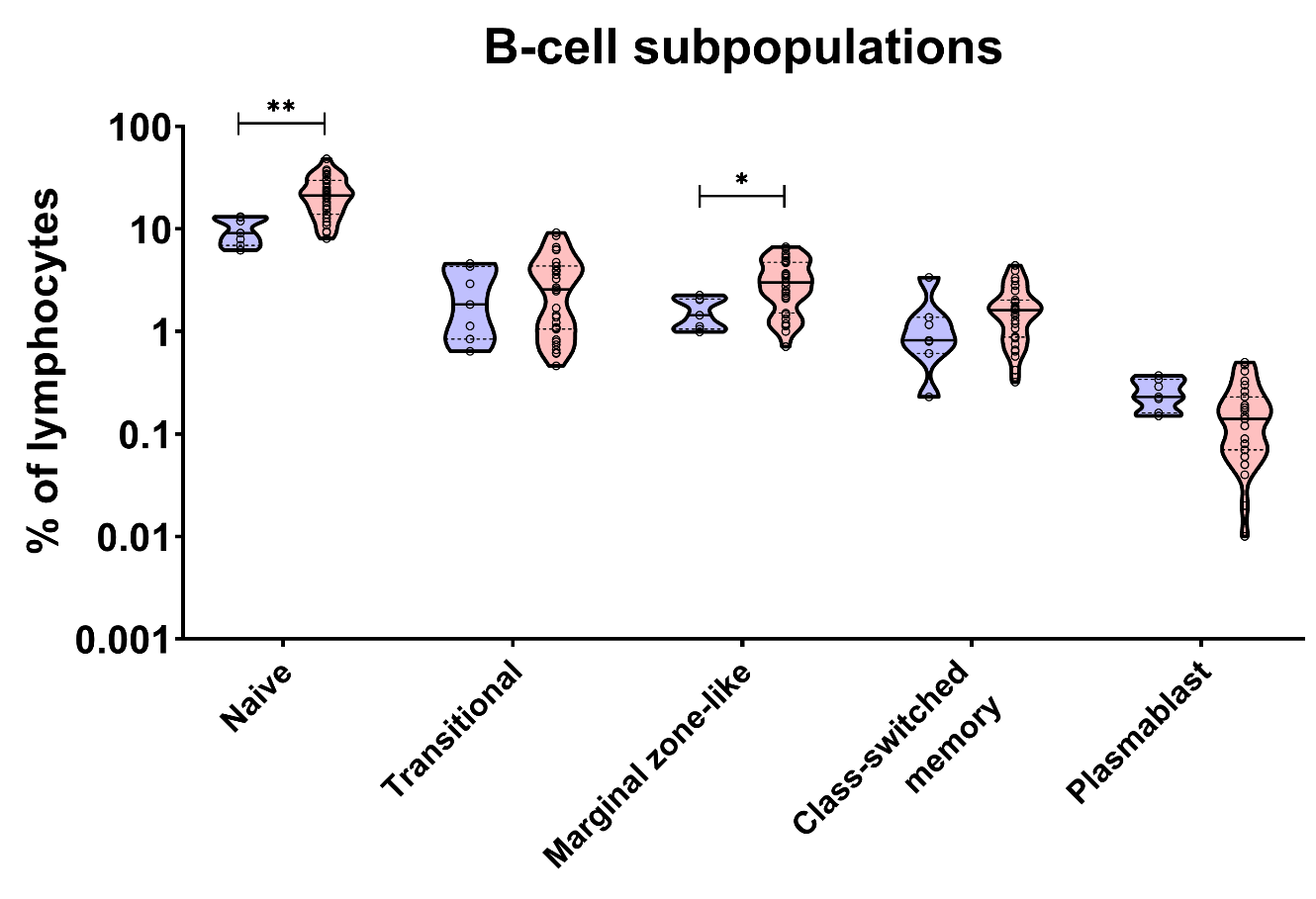
