## Supplementary material for "B cells, BAFF and interferons in MIS-C": Table 1

Table 1 Description of cohorts. MIS-C = Multisystem Inflammatory Syndrome in Children associated with COVID-19; HD = healthy donor; COVID = coronavirus disease; SD = standard deviation.

|  | **n** | **Sex** | **Age**  **years range**  **(mean ± SD)** |
| --- | --- | --- | --- |
| MIS-C | 50 | 24 female | 0.9-18 (7.8 ± 4.35) |
| MIS-C convalescent | 4 | 2 female | 1.4-5.5 (3.5 ± 1.4) |
| HD children post-COVID | 7 | 2 female | 1-14 (9.9 ± 3.9) |
| HD children | 8 | 5 female | 11.6-17.2 (13.7 ± 1.9) |
